# BASILICA or snorkel stenting to prevent or treat coronary obstruction: Multicenter international COBRA-TAVR registry

**DOI:** 10.1101/2025.05.12.25327191

**Authors:** Hiroki A. Ueyama, John C. Lisko, Adam B. Greenbaum, Isida Byku, Patrick T. Gleason, Chandan M. Devireddy, Norihiko Kamioka, Toby Rogers, Jaffar M. Khan, Gaetano Paone, Robert J. Lederman, Vasilis C. Babaliaros, COBRA Investigators

## Abstract

Coronary obstruction is a rare but life-threatening complication following transcatheter aortic valve replacement (TAVR). Comparative analysis between snorkel stenting and BASILICA (Bioprosthetic or native Aortic Scallop Intentional Laceration to prevent Iatrogenic Coronary Artery obstruction) remain limited. We analyzed 122 patients from the COBRA registry, including 68 who underwent BASILICA and 54 who received snorkel stents. In-hospital survival was numerically higher in the BASILICA group compared to the snorkel stent group (97% vs. 89%). BASILICA was also associated with lower rates of life-threatening bleeding (0.0% vs. 10%) and major vascular complications (1.5% vs. 12%). In patients at risk, a preemptive BASILICA strategy may offer an attractive approach for primary prevention of coronary obstruction in patients undergoing TAVR.

## Text

Coronary obstruction following transcatheter aortic valve replacement (TAVR) occurs when the transcatheter heart valve displaces the native or bioprosthetic valve leaflets into an “open” position—occluding the coronary ostia or Sinus of Valsalva. While the incidence is low, this complication portends a high mortality. Available techniques to treat or prevent coronary obstruction are prepositioning a coronary “snorkel” stent or BASILICA (electrosurgical <u>B</u>ioprosthetic or native <u>A</u>ortic <u>S</u>callop Intentional <u>L</u>aceration to prevent Iatrogenic <u>C</u>oronary <u>A</u>rtery obstruction). (1,2) While outcomes of both techniques have been reported in single-arm multicenter trials or registries, there are no published comparisons. Herein, we report in-hospital clinical outcomes of patients undergoing either technique to prevent or treat coronary obstruction after TAVR.

The cohort was derived from an investigator-initiated, multicenter, international registry (COBRA) including patients undergoing TAVR with high risk of coronary obstruction between 2008 to 2020 (BASILICA: 2017 to 2020; snorkel stenting: 2008 to 2019). Patients were deemed to be at high risk by the enrolling site and underwent either preventive/ therapeutic “snorkel” stent or BASILICA. (1,2) Clinical outcomes were compared at discharge and defined by Valve Academic Research Consortium-2 definitions (contemporaneous to the analysis period).(3) Measures of statistical significance are not provided because of the small sample size. The Emory University Institutional Review Board approved this study, and the participating sites either obtained local IRB approval or accepted the Emory IRB exemption.

A total of 122 patients from 9 centers in North America, Europe, and Japan were included (68 BASILICA and 54 snorkel stent) (Figure 1). Patients were old (age 78.3 ± 8.9) with high surgical risk (STS PROM score: BASILICA 7.2% ± 5.9 vs. snorkel stent 6.6% ± 4.4). A balloon expandable valve was used in 80% of cases. Most underwent transfemoral access (82% BASILICA, 96% snorkel stent); 18% underwent transcaval access in BASILICA and 3.7% underwent transapical access in snorkel stent cohort. Snorkel stent was performed prophylactically in 43% and as bailout after obstruction in 57% of cases. Most snorkel stenting addressed single vessel obstruction (98%), compared with the BASILICA cohort where double BASILICA was performed in 47%. Two patients (3.0%) required placement of an orthotopic stent for leaflet prolapse after BASILICA.

**Figure 1:**
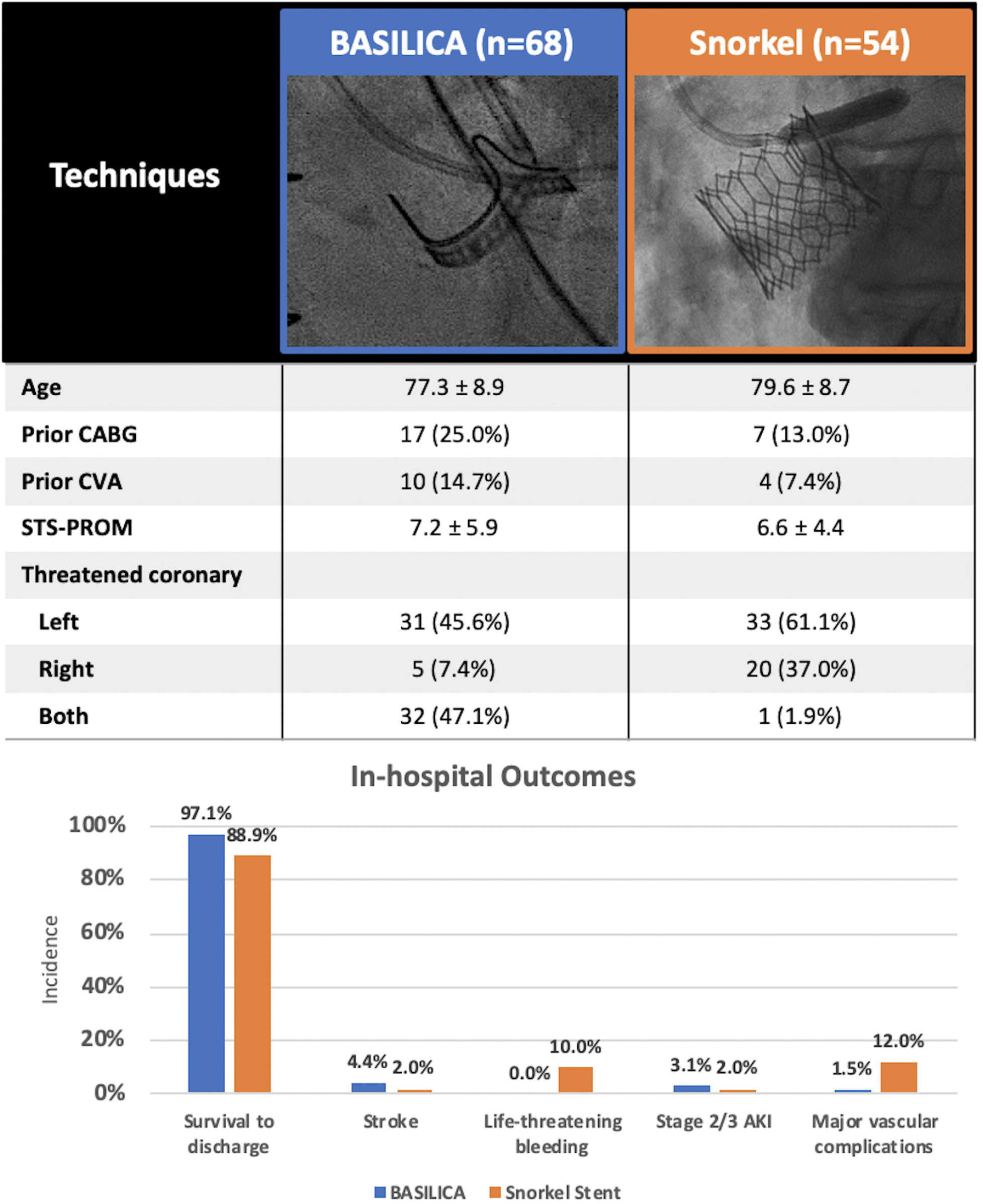
Patient characteristics and in-hospital outcomes AKI = acute kidney injury, CABG = Coronary artery bypass grafting, CVA = cerebrovascular accident, STS-PROM = Society of Thoracic Surgeons Predicted Risk of Mortality

In-hospital survival was numerically higher in the BASILICA cohort compared to snorkel stenting (97% vs. 89%, respectively). All-stroke occurred in 4.4% of patients undergoing BASILICA compared with 2.0% undergoing the snorkel stent. The rates of life-threatening bleeding (0.0% vs. 10%) and major vascular complications (1.5% vs. 12%) were numerically lower in BASILICA compared with snorkel stenting. Results were unchanged when excluding transapical transapical access.

BASILICA is attractive in that it is a primary prevention strategy with infrequent failure requiring bailout stent (3% in this series), it facilitates re-access of threatened coronary arteries, and it allows bailout with orthotopic stenting. Disadvantages of BASILICA include that it requires multiple procedure steps. There are conflicting suggestions of excess stroke.(1,4)

Snorkel stenting is attractive in its simplicity. Disadvantages include crushing of the stent between the transcatheter heart valve and aorta, the risk of subacute and late stent thrombosis, and difficulty of re-engaging the threatened coronary artery, which may limit treatment of both stable and acute coronary syndromes. Data on long-term outcomes after snorkel stenting are absent.

There are several limitations to this study. First, this was a retrospective study with self-reported data. The data are neither monitored nor audited. The study was not powered to detect significant differences in outcomes. A number of patients in the snorkel stent cohort had the procedure done as a bailout strategy after coronary obstruction and subsequent myocardial injury may have already occurred; this may partially explain the higher mortality rate in this group. The use of cerebral protection during TAVR was not systematically collected in this dataset to explain the rate of stroke. Finally, it is unclear as to why snorkel stent was associated with high rate of bleeding and vascular complications, but we suspect it is related to the emergency use of mechanical support as a bailout.

In conclusion, a preemptive strategy with the use of BASILICA to prevent coronary obstruction had numerically higher survival and fewer adverse events compared with preemptive or bailout snorkel stenting.

## Data Availability

All data produced in the present study are available upon reasonable request to the authors

## Acknowledgement

*We thank Dr. Sebastian Ludwig from University Heart and Vascular Center Hamburg, Hamburg, Germany; Dr. Raj Makkar and Dr. Sung-Han Yoon from Cedars Sinai Medical Center, California, USA; Dr. Tomohiro Kawaguchi and Dr. Sinichi Shirai from Kokura Memorial Hospital, Kitakyushu Japan; Dr. Masanori Yamamoto and Dr. Ai Kagase from Toyohashi Heart Center/Nagoya Heart Center, Nagoya, Japan; Dr. Norio Tada and Dr. Masaki Miyasaka from Sendai Kosei Hospital, Sendai, Japan; Dr. Masahiro Yamawaki and Dr. Yosuke Honda from Saiseikai Yokohama Tobu Hospital, Yokohama, Japan; and Dr.Giuseppe Muscogiuri, Dr. Laura Fusini, and Dr. Gianluca Pontone from Centro Cardiologico Monzino IRCCS, Milan, Italy for their significant contribution to the multicenter study.

## Abbreviations

BASILICA: Bioprosthetic or native aortic scallop intentional laceration to prevent iatrogenic coronary artery obstruction
STS-PROM: Society of Thoracic Surgeons Predicted Risk of Mortality
TAVR: transcatheter aortic valve replacement

## Notes

**Funding:** Supported by Emory Division of Cardiology intramural funds and by NIH Z01-HL006040.

**Conflict of Interest/Financial Disclosures:** ABG and VCB have received institutional research support from Abbott Vascular, Ancora Heart, Edwards Lifesciences, Gore Medical, JenaValve, Medtronic, Polares Medical, Transmural Systems, and 4C Medical; have received consulting fees from Abbott Vascular, Edwards Lifesciences, and Medtronic; and have an equity interest in Transmural Systems. IB and PTG have institutional research contracts for clinical investigation of transcatheter aortic, mitral, and tricuspid devices from Edwards Lifesciences, Abbott Vascular, Medtronic, and Boston Scientific. CMD receives compensation for the following roles: Medtronic: Consultant; Edwards Lifesciences: Proctor; ReCor Medical: Consultant; and Shockwave Medical: Consultant. NK has served as a proctor for Edwards Lifesciences. TR has served as a proctor and consultant for Boston Scientific, Edwards Lifesciences, and Medtronic; has served on the advisory board for Medtronic; and holds equity interest in Transmural Systems Inc. GP has institutional research contracts for clinical investigation of transcatheter aortic, mitral, and tricuspid devices from Edwards Lifesciences, Abbott Vascular, Medtronic, and Boston Scientific; and is a proctor for Edwards Lifesciences. RJL is coinventor on patents assigned to the National Institutes of Health (NIH), for electrosurgical devices. All other authors have reported that they have no relationships relevant to the contents of this paper to disclose.

### Competing Interest Statement

ABG and VCB have received institutional research support from Abbott Vascular, Ancora Heart, Edwards Lifesciences, Gore Medical, JenaValve, Medtronic, Polares Medical, Transmural Systems, and 4C Medical; have received consulting fees from Abbott Vascular, Edwards Lifesciences, and Medtronic; and have an equity interest in Transmural Systems. IB and PTG have institutional research contracts for clinical investigation of transcatheter aortic, mitral, and tricuspid devices from Edwards Lifesciences, Abbott Vascular, Medtronic, and Boston Scientific. CMD receives compensation for the following roles: Medtronic: Consultant; Edwards Lifesciences: Proctor; ReCor Medical: Consultant; and Shockwave Medical: Consultant. NK has served as a proctor for Edwards Lifesciences. TR has served as a proctor and consultant for Boston Scientific, Edwards Lifesciences, and Medtronic; has served on the advisory board for Medtronic; and holds equity interest in Transmural Systems Inc. GP has institutional research contracts for clinical investigation of transcatheter aortic, mitral, and tricuspid devices from Edwards Lifesciences, Abbott Vascular, Medtronic, and Boston Scientific; and is a proctor for Edwards Lifesciences. RJL is coinventor on patents assigned to the National Institutes of Health (NIH), for electrosurgical devices. All other authors have reported that they have no relationships relevant to the contents of this paper to disclose.

### Funding Statement

Supported by Emory Division of Cardiology intramural funds and by NIH Z01-HL006040.

### Author Declarations

The Emory University Institutional Review Board approved this study, and the participating sites either obtained local IRB approval or accepted the Emory IRB exemption

## References

1. Khan JM, Babaliaros VC, Greenbaum AB et al. Preventing Coronary Obstruction During Transcatheter Aortic Valve Replacement: Results From the Multicenter International BASILICA Registry. JACC Cardiovasc Interv 2021;14:941–948.

2. Mercanti F, Rosseel L, Neylon A et al. Chimney Stenting for Coronary Occlusion During TAVR. JACC: Cardiovascular Interventions 2020;13:751–761.

3. Kappetein AP, Head SJ, Généreux P et al. Updated Standardized Endpoint Definitions for Transcatheter Aortic Valve Implantation. Journal of the American College of Cardiology 2012;60:1438–1454.

4. Khan JM, Greenbaum AB, Babaliaros VC et al. The BASILICA Trial: Prospective Multicenter Investigation of Intentional Leaflet Laceration to Prevent TAVR Coronary Obstruction. JACC Cardiovasc Interv 2019;12:1240–1252.

